# AI-MI: A Deep Learning Model to Predict Actionable Acute Coronary Syndrome Using 12-Lead ECGs

**DOI:** 10.1101/2025.05.18.24319528

**Authors:** Yuri Ahuja, Prabhu Sasankan, Robert Ronan, Larry Chinitz, Sunil Rao, Lior Jankelson

## Abstract

**Background:** Chest pain is among the most common chief complaints in Emergency Departments (EDs), and differentiating acute coronary syndrome from low-risk chest pain etiologies is often nontrivial. Early and accurate identification of acute coronary syndrome (ACS) is a key quality metric in Emergency Departments as diagnostic delay results in myocardial infarction, while unnecessary coronary angiography leads to adverse effects, prolonged hospital stays, and poor utilization of cardiac catheterization laboratory resources.

**Objectives:** We present AI-MI, a novel deep learning technology that predicts whether a patient presenting with chest pain will undergo same-encounter coronary revascularization.

**Methods:** We use a 1D ResNet convolutional neural network architecture pretrained on an arrythmia- detection task as a foundation model, and we train it using Electronic Health Record data from NYU Langone Health EDs from 2009 to 2024. We define three cohorts within this dataset – patients from whom a troponin has been ordered, those with a positive troponin, and those scheduled for coronary angiogram – to evaluate our model’s applicability at sequential stages in the ACS diagnostic/therapeutic algorithm.

**Results:** AI-MI achieves AUROCs of 0.91, 0.81, and 0.67, and AUPRCs of 0.35, 0.39, and 0.57, within the “troponin ordered,” “troponin positive,” and “coronary angiogram” cohorts respectively. AI- MI trained using sequential ECGs consistently and significantly outperformed that using individual ECGs.

**Conclusions:** AI-MI can be used in the point-of-care evaluation of chest pain to improve differentiation of ACS from low-risk chest pain, both promoting early cardiology consultation for revascularization and decreasing the rate of unnecessary coronary angiography.

## Introduction

Chest pain is the second leading cause of emergency department (ED) visits for adults in the United States, with nearly 11 million annual chest pain encounters accounting for ∼5.5% of all ED visits.^1^ Only ∼5.1% of chest pain cases in the ED have acute coronary syndrome (ACS), while more than half will ultimately be diagnosed with noncardiac etiologies.^2^ However, promptly identifying ACS patients is vital in ensuring timely coronary revascularization, while reliably ruling out low-risk patients is crucial to optimizing allocation of limited cardiac catheterization laboratory resources, enabling timely discharge, avoiding unnecessary hospital admissions, and avoiding the adverse effects of unnecessary coronary angiography.^3–5^

Currently, risk stratification of chest pain relies on patient history, physical examination, serial electrocardiograms (ECGs), and cardiac biomarker (i.e. troponin) evaluation to distinguish low-risk chest pain from ACS.^6^ However, this approach is limited by interobserver variability, subtlety of ECG changes (particularly early in ischemia), and the time lag in biomarker elevation, exposing patients to morbidity both from delayed revascularization in true positive cases and unnecessary invasive angiography in false positives cases. Distinguishing type 1 myocardial infarction (MI) from type 2 MI, which is unlikely to benefit from revascularization, remains particularly challenging due to overlapping clinical presentations and biomarker profiles, leading to frequent reliance on invasive coronary angiography to make this distinction.^7^

Artificial intelligence (AI) and deep learning have shown considerable potential in enhancing cardiovascular diagnostics, particularly in the detection of arrhythmias and prediction of structural heart diseases from standard 12-lead ECG waveforms.^8^ Recently, interest has grown in the use of AI models to predict myocardial infarction from 12-lead ECGs, with models demonstrating strong performance in predicting a discharge diagnosis of MI^9^ or the presence of occlusive myocardial infarction in patients presenting with chest pain.^10^ Despite these advancements, a gap remains in the application of AI to predict whether a patient with anginal symptoms will ultimately undergo coronary revascularization, a highly desirable predictive task with direct relevance in clinical management.

In this study, we introduce a novel deep learning approach, AI-MI, that uses serial ECG waveforms to predict whether chest pain patients being evaluated for ACS ultimately undergo coronary revascularization within the same hospital encounter. We train and evaluate AI-MI in three cohorts of patients representing sequential stages in the ACS workup: patients with a troponin assay ordered, a positive troponin assay, and a scheduled coronary angiogram. We additionally evaluate whether models trained on sequential ECG tracings demonstrate improved performance over models trained on single 12-lead ECG tracings, which is the standard approach in the literature.

## Methods

### Data Source

We collected Electronic Health Record data between 2012 and 2022 from all Emergency Department (ED) encounters at the NYU Langone Health system in New York, NY. NYU Langone Health includes 5 adult hospitals in the New York area: Tisch Hospital, Kimmel Pavilion, NYU Langone Orthopedic Hospital, NYU Langone Hospital – Brooklyn, and NYU Langone Hospital – Long Island. Each ED encounter encompasses the period between ED presentation and ED or hospital discharge. To select for encounters wherein acute coronary syndrome is on the differential diagnosis, we selected only ED encounters wherein a patient had1) at least one 12-lead ECG performed, and 2) at least one troponin assay ordered. Apart from ECG waveforms, we collected results for all troponin assays (troponin-T, troponin-I, high sensitivity troponin) and CPT procedure codes for coronary angiogram, percutaneous coronary intervention (PCI), and coronary artery bypass graft (CABG). The NYU Langone Institutional Review Board (IRB) reviewed and approved our use of NYU Langone patient data for the purposes of this study.

### Cohorts and Outcomes

We defined three cohorts of interest in this study. The first cohort consists of all patients with anginal symptoms for whom a troponin assay has been ordered but is not necessarily positive (henceforth the “troponin ordered” cohort). The second cohort consists of all patients with anginal symptoms for whom a troponin assay has resulted positive, defined as a troponin-T >= 0.05 ug/mL, a troponin-I >= 0.05 µg/mL, or a troponin-HS >= 5 µg/mL (henceforth the “troponin positive” cohort). The third cohort consists of all patients with anginal symptoms for whom a coronary angiogram was performed. This cohort includes patients deemed by an interventional cardiologist to be at sufficient risk of ACS to warrant invasive coronary angiography. In our dataset, 123,397 encounters met criteria for cohort 1, 32,163 for cohort 2, and 19,716 for cohort 3.

We defined our outcome of interest as the binary indicator of whether a patient underwent coronary revascularization – namely, percutaneous coronary intervention (PCI) or coronary artery bypass grafting (CABG) – during an index encounter.

### ECG Data Preprocessing

ECG waveforms were downsampled from 500 Hz to 250 Hz for 10 second periods, for a total of 2500 timepoints. 12-lead ECGs were thus represented as (12, 2500) dimensional arrays. Various waveform filter and data normalization strategies were considered, but no such data pre- processing techniques were utilized as they were found to degrade model performance.

We experimented with prediction using individual and sequential ECG scans. For the latter, sequential ECGs within a patient encounter were each truncated to the first 2048 timepoints and then concatenated to produce the model input. In other terms, given two sequential ECGs of dimension (12, 2500), each was truncated to dimension (12, 2048) and then concatenated to create a (12, 4096) dimensional array.

### Model Selection and Training

We experimented with multiple 1D CNN and vision transformer foundation models, including ResNet1D,^11^ EfficientNet,^12^ ConvNet,^13^ ConvNext,^14^ ViT,^15^ BeIT,^16^ DeIT,^17^ and Swin.^18^ Ultimately, we found that a 1D ResNet CNN pretrained to identify arrhythmias on ECGs from the Massachusetts Institute of Technology – Beth Israel Hospital (MIT-BIH) Arrhythmia Database maximized out-of-sample AUROC when trained and validated on our dataset for same- encounter revascularization, so we used this foundation model to develop AI-MI.^19^ We removed the final classification layer of the above foundation model, and in its place included a 64- dimensional dense hidden layer followed by a 1-dimensional binary classifier.

We trained AI-MI using an Adam optimizer in Python’s Keras package. We enabled learning rate reduction on plateau with factor 0.2, patience 5, and minimum learning rate 1e-6. Moreover, we enabled early stopping with patience 20 and minimum delta 1e-5. We ran validation experiments to optimize the starting learning rate, maximum number of epochs, and batch size, ultimately setting these hyperparameters to 1e-3, 50, and 32 respectively.

### Metrics and Analyses

To evaluate AI-MI, we estimated area under the receiver operating characteristic curve (AUROC), area under the precision recall curve (AUPRC), F1 score, sensitivity, specificity, positive predictive value (i.e. precision), and negative predictive value. Confidence intervals around AUROC and AUPRC estimators were obtained by bootstrapping with 100 subsamples.

We implemented the Grad-CAM algorithm to visually probe the features that drive AI-MI’s predictions.^20^ We applied Grad-CAM to the “troponin ordered” cohort.

### Thresholding

We discretize our probability predictions using two thresholds – one to achieve high (95%) sensitivity (to be used as a “rule-out” test) and one to achieve high specificity (to be used as a “rule-in” test). The specificity threshold was set to 95%, 75%, and 50% for the “troponin ordered,” “troponin positive”, and “coronary angiogram” cohorts. Thresholds were ascertained using a held-out validation set separate from the test set used to measure model performance. We envision predictions below the lower (95% sensitivity) threshold to be interpreted as negative for ACS, those above the upper (high specificity) threshold to be interpreted as positive for ACS, and those between the thresholds to be interpreted as indeterminate. A schematic of this approach is displayed in **Figure 1**.

**Figure 1:**
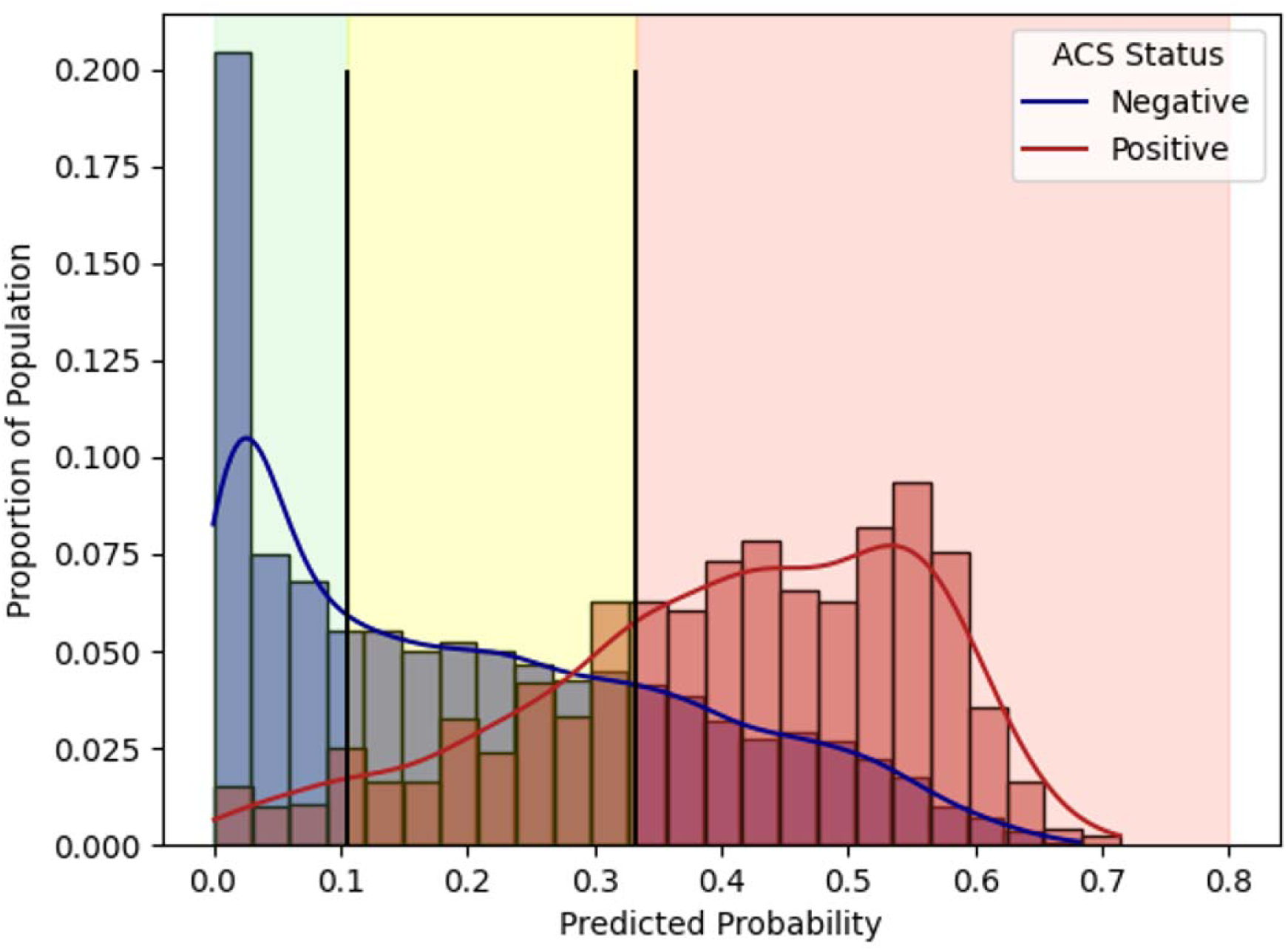
Schematic of our proposed low-threshold/high-threshold risk predictor. Cases with predicted scores below the lower threshold are taken as negative, and those with predicted scores above the higher threshold are taken as positive. Cases with predicted scores between the thresholds are indeterminate. Lower thresholds are set to achieve 95% sensitivity; higher thresholds are set to achieve a 95%, 75%, and 50% specificity in our troponin ordered, troponin positive, and coronary angiogram ordered datasets respectively.

### Software

We used Python 3.9.16 with packages numpy v1.21.0,^21^ scipy v1.13.1,^22^ pandas v2.0.2,^23^ scikit- learn v1.5.1,^24^ tensorflow v2.12.0,^25^ and keras v2.12.0.^26^ We used keras to train and deploy our deep learning models.

## Results

### Study Population

Of the 123,397 patient encounters in the “troponin ordered” cohort, 11.1% underwent coronary angiogram and 4.5% underwent coronary revascularization. Of the 32,163 patient encounters in the “troponin positive” cohort, 30.9% underwent coronary angiogram and 14.2% underwent coronary revascularization. Of the 19,716 patient encounters in the “coronary angiogram” cohort, 44.4% underwent coronary revascularization. Mean demographics of the three study cohorts are outlined in **Table 1**.

**Table 1:** Demographics and statistics for our three cohorts of interest: 1) ED patients for whom a troponin was ordered, 2) ED patients for whom a troponin resulted positive, and 3) ED patients sent for urgent coronary angiography.

|  | Troponin Ordered | Troponin Positive | Coronary Angiogram |
| --- | --- | --- | --- |
| **Number of encounters** | 123,397 | 32,163 | 19,716 |
| **PCI/CABG Prevalence** | 4.5% | 14.2% | 44.4% |
| **Cath/CABG Prevalence** | 11.1% | 30.9% | 100.0% |
| **Mean Age** | 55.5 | 65.4 | 65.9 |
| **Percent Male** | 50.9% | 60.1% | 68.9% |
| **Percent Caucasian** | 56.2% | 60.0% | 66.2% |
| **Percent African American** | 15.1% | 14.3% | 8.1% |
| **Percent Asian** | 1.7% | 2.0% | 1.9% |
| **Percent Native American** | 0.1% | 0.2% | 0.1% |
| **Percent Other** | 26.9% | 23.5% | 23.7% |

### Prediction of Same-Encounter Coronary Revascularization

As displayed in **Figure 2**, our algorithm achieved AUROCs of 0.91, 0.81, and 0.67, and AUPRCs of 0.35, 0.39, and 0.57, within the “troponin ordered”, “troponin positive”, and “coronary angiogram” cohorts respectively.

**Figure 2:**
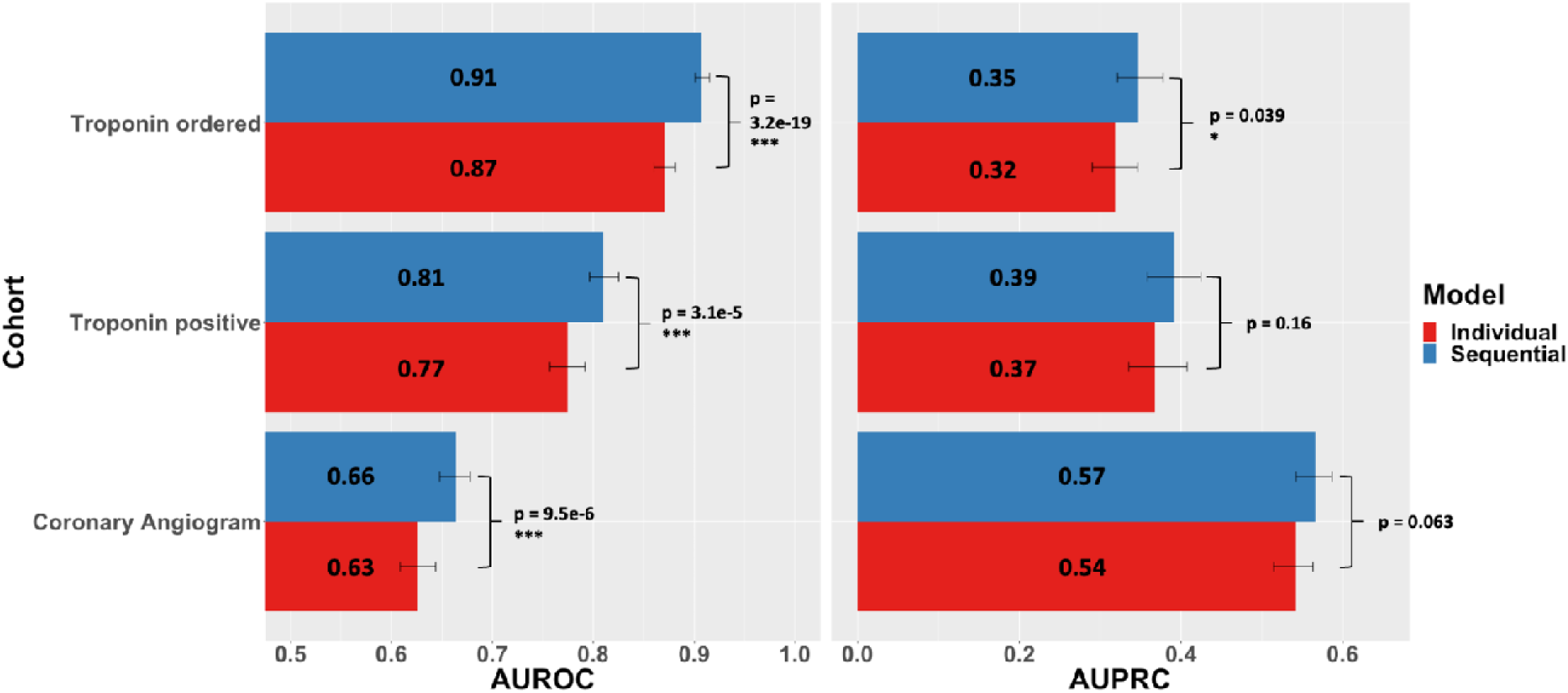
Areas under the receiver operating curve (left) and precision recall curve (right) for machine learning models trained on individual ECG scans (red) versus sequential scans (blue). AI-MI is the first of its kind that is trained using sequential rather than individual ECGs, enabling it to capture the rich information inherent to evolutions in a patient’s ECG over time.

Within the “troponin ordered” cohort, AI-MI achieved 95.2% sensitivity and 99.6% negative predictive value at the lower threshold, and 95.3% specificity and 35.5% positive predictive value at the upper threshold. Within the “troponin positive” cohort, our algorithm achieved 91.4% sensitivity and 96.6% negative predictive value at the lower threshold, and 76.3% specificity and 34.2% positive predictive value at the upper threshold. Within the “coronary angiography” cohort, AI-MI achieved 94.3% sensitivity and 81.7% negative predictive value at the lower threshold, and 49.5% specificity and 50.5% positive predictive value at the upper threshold. Detailed results can be found in **Table 2**, and confusion matrices can be found in **Supplementary Tables S1-S4**. Receiver operating characteristic and precision recall curves are displayed in **Figure 3**. Removing STEMI cases – which tend to be easily identifiable – from the cohort did not meaningfully affect these results, as demonstrated in **Supplementary Figure S1**.

**Figure 3:**
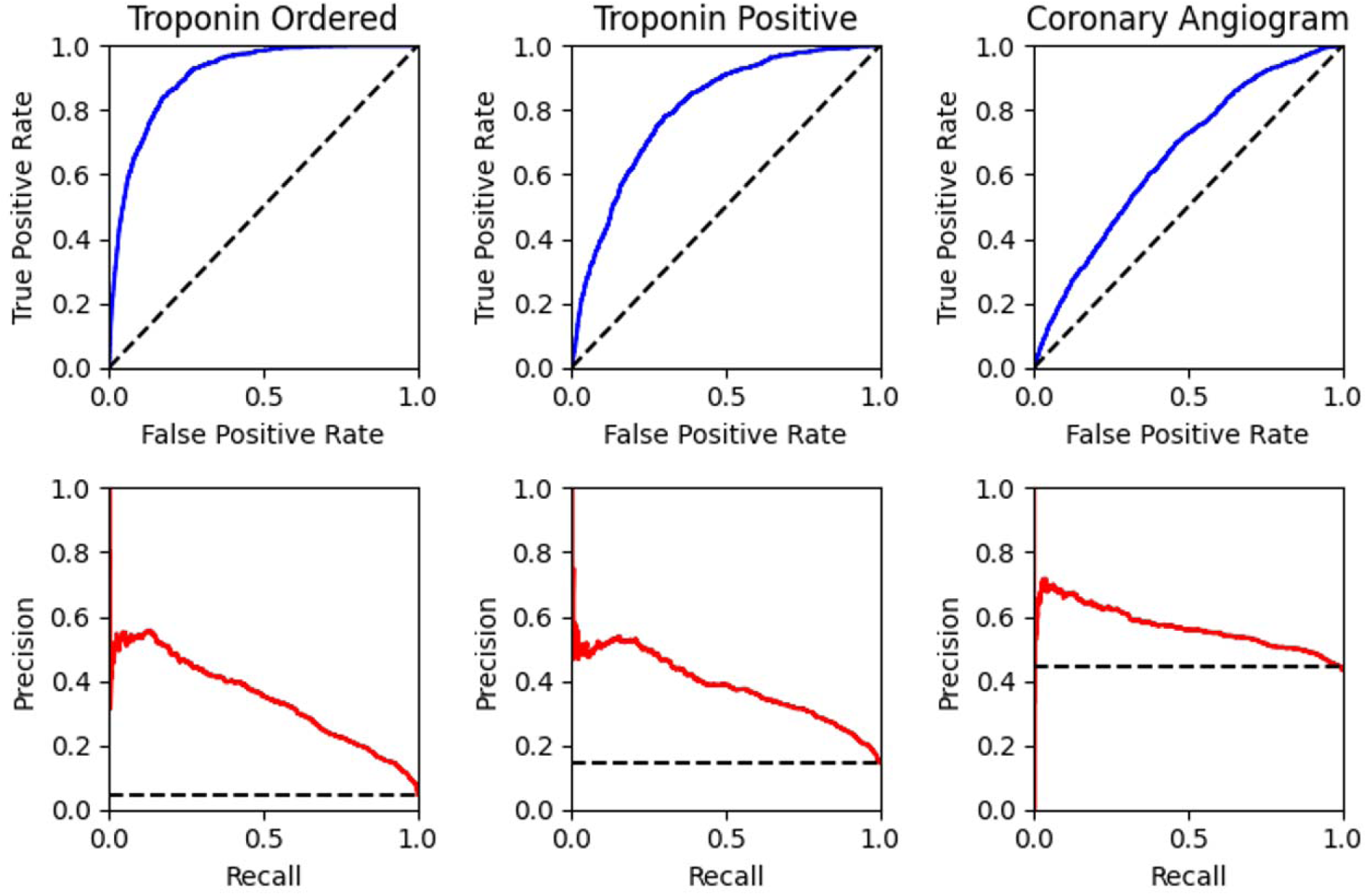
Areas under the receiver operating curve (top, blue) and precision recall curve (bottom, red) for AI-MI within cohorts for whom a troponin assay has been ordered (left), a troponin assay has resulted positive (middle), and a coronary angiogram has been scheduled (right). Black dotted lines represent random predictors (i.e. AUROC = 0.5, AUPRC = outcome prevalence).

**Table 2:** Key metrics of AI-MI predicting coronary revascularization during an index admission among ED patients with anginal symptoms. Lower (i.e. high-sensitivity) thresholds are set for the “troponin ordered,” “troponin positive,” and “coronary angiogram” cohorts such that AI-MI achieves 95% sensitivity in held-out validation sets for each cohort, enabling its use to “rule-out” ACS. Upper (i.e. high-specificity) thresholds are set such that AI-MI achieves 95%, 75%, and 50% specificity in held-out validation sets of the above cohorts respectively, enabling its use to “rule-in” ACS.

| Cohort | Threshold | Sensitivity | Specificity | PPV | NPV | F1 Score |
| --- | --- | --- | --- | --- | --- | --- |
| **Troponin ordered** | Lower | 0.952 | 0.656 | 0.122 | 0.996 | 0.216 |
|  | Upper | 0.514 | 0.953 | 0.355 | 0.975 | 0.420 |
| **Troponin positive** | Lower | 0.914 | 0.442 | 0.226 | 0.966 | 0.362 |
|  | Upper | 0.693 | 0.763 | 0.342 | 0.933 | 0.459 |
| **Coronary angiogram** | Lower | 0.943 | 0.184 | 0.456 | 0.817 | 0.615 |
|  | Upper | 0.710 | 0.495 | 0.505 | 0.702 | 0.590 |

### Sequential versus Individual ECG Models

AI-MI is the first of its kind to predict using a concatenation of sequential ECG scans rather than just individual scans, enabling it to capture the rich information present in the evolution of ECG parameters over time. To study the benefit of using sequential scans, we compared AI-MI to a pretrained model using individual ECGs only. Predicting coronary revascularization using sequential EKGs significantly outperformed using individual EKGs, with an increase in AUROC from 0.87 to 0.91, 0.77 to 0.81, and 0.63 to 0.66 for the troponin ordered, troponin positive, and coronary angiogram groups respectively. Sequential prediction achieved a significantly higher AUPRC within the “troponin ordered” cohort but was statistically equivalent in other cohorts. Mean AUROCs and AUPRCs are displayed in **Figure 2**.

### Using Grad-CAM to Interrogate Predictions

We applied the Grad-CAM algorithm to obtain an intuitive visual understanding of what components of the ECG drive AI-MI’s predictions. **Figure 4** depicts the Grad-CAM algorithm applied to two ECGs within the “troponin ordered” cohort: AI-MI’s highest probability prediction that turned out to be true (“top true positive”) and AI-MI’s highest probability prediction that turned out to be false (“top false positive”). ECGs with the next nine highest probabilities (true and false positives respectively) are included in **Supplementary Figure S2**. AI-MI appears to focus primarily on the PR and ST segments, followed by the QRS complex and T wave.

**Figure 4:**
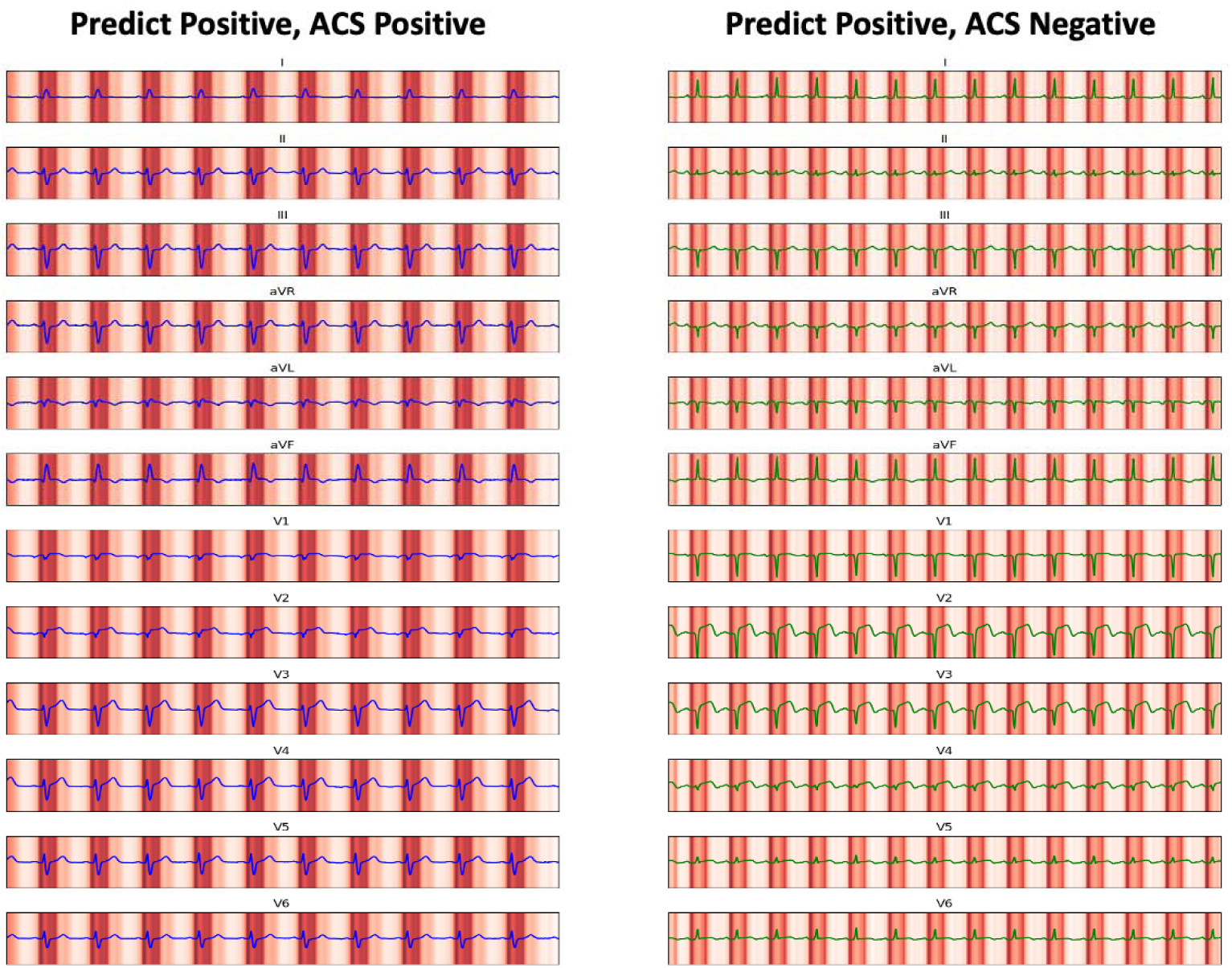
Heatmap displaying regions of ECGs driving AI-MI’s predictions for (left) the patient with ACS for whom AI-MI predicted the highest probability of ACS (i.e. top true positive), and (right) the patient without ACS for whom AI-MI predicted the highest probability of ACS (i.e. top false positive). Heatmaps were generated using the Grad-CAM algorithm. Heatmaps for the next nine top true positives and false positives are displayed in **Supplementary Figure S2**.

### Predicting Readmission for Revascularization

AI-MI’s predicted probability of same-encounter revascularization has an AUROC of 0.680 and an AUPRC of 0.105 when used to predict 30-day re-admission for revascularization among patients discharged without having received a coronary angiogram. Detailed results are shown in **Table 3**. Examples of initial EKGs during patients’ index admission and re-admission for revascularization are shown in **Supplementary Figure S3**.

**Table 3:** Metrics using AI-MI’s predictions to predict re-admission for ACS and revascularization among patients discharged without coronary angiography. Thresholds are set to achieve 95% sensitivity (lower) and 95% specificity (upper).

| Threshold | AUROC | AUPRC | Sensitivity | Specificity | PPV | NPV | F1 Score |
| --- | --- | --- | --- | --- | --- | --- | --- |
| **Low** | 0.680 | 0.105 | 0.773 | 0.385 | 0.065 | 0.968 | 0.121 |
| **High** | 0.680 | 0.105 | 0.409 | 0.808 | 0.106 | 0.961 | 0.168 |

## Discussion

In patients with anginal symptoms in the ED, the early identification of ACS is critical to reduce morbidity and mortality and is a key quality metric of emergency departments and hospital wards alike. However, variable clinical presentation and ECG changes in ACS – particularly in unstable angina (UA)/non-ST elevation myocardial infarction (NSTEMI) – are often subtle or nonspecific, limiting human inference on the presence of an occluded coronary vessel as the cause of symptoms **(Figure 5**). This work aims to utilize recent developments in AI to augment the ECG for decision support in patients with anginal symptoms.

**Figure 5:**
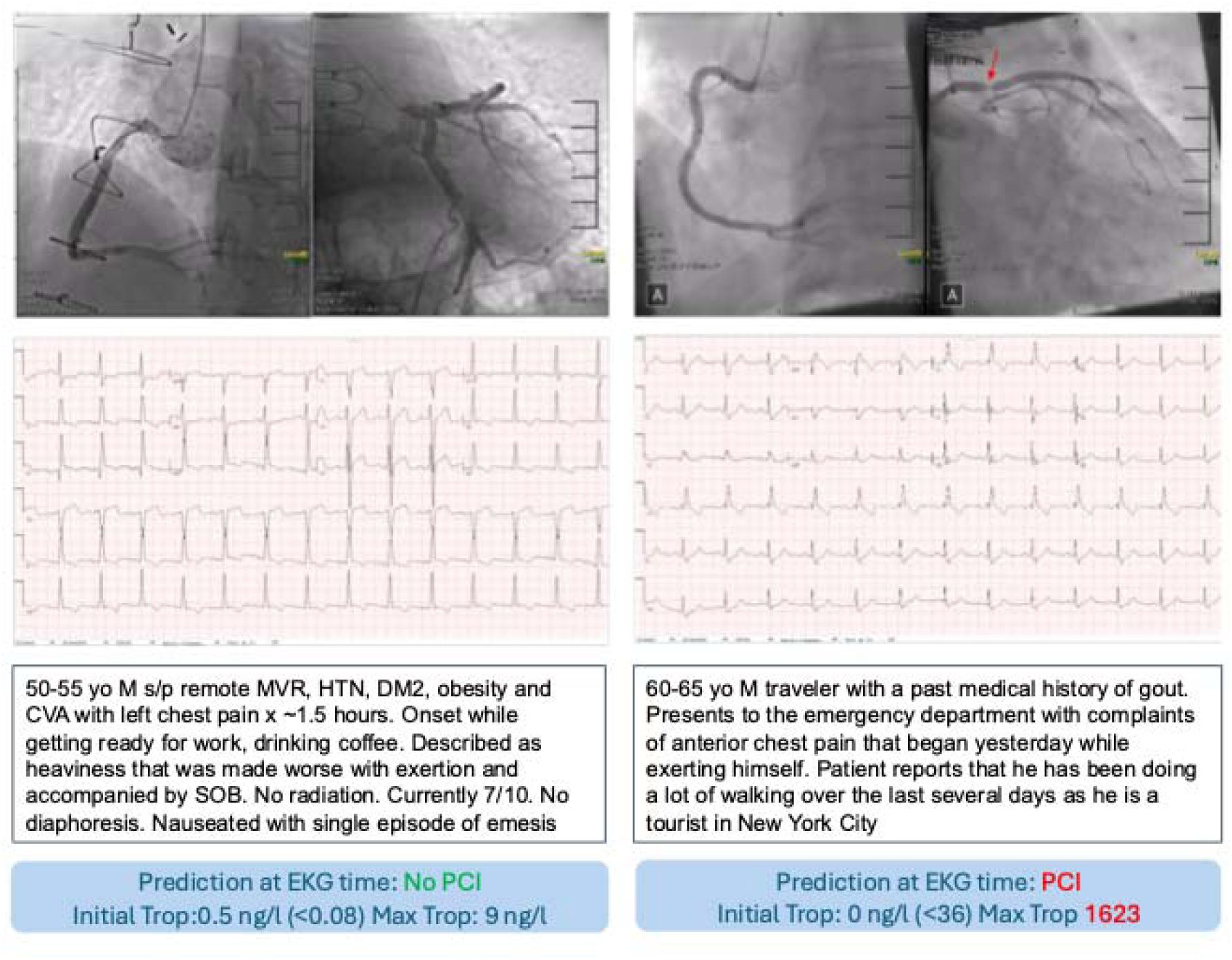
Examples of clinically similar chest pain cases for which AI-MI predicted “No PCI” and the patient did not have significant coronary obstruction on coronary angiogram (left), and for which AI-MI predicted “PCI” and the patient did have significant coronary obstruction (right). Historical details and visually evident ECG features are often insufficient to differentiate ACS, and troponin elevations can lag hours behind the onset of ischemia.

AI-MI achieved high discriminatory accuracy in the direct task of predicting the need for a coronary intervention, enabling its use to augment provider decision making at the point of care. We chose this outcome, rather than the diagnosis of occlusion myocardial infarction (OMI) – to get as close as possible to the most impactful action item in the ACS therapeutic algorithm.

Given the critical importance of the cardiac catheterization laboratory as a limited resource and arate limiting step in the resolution of an ACS, we suggest utilization of our algorithm as a decision support aid in the prioritization of patients for invasive management.

AI-MI’s predictive performance within the “troponin ordered” cohort demonstrates its ability to identify ACS among ED patients with anginal symptoms, enabling *early* cardiology consultations while preventing *unnecessary* consultations. Its performance in the “coronary angiogram” cohort reflects an improvement on the clinical judgment of an interventional cardiologist who has already opted for angiography, possibly influencing the decision to proceed with angiography and/or influencing which patients should be prioritized for limited cardiac catheterization laboratory resources. Finally, our analysis of 30-day readmission for revascularization suggests that AI-MI’s predictions may be used to prevent inappropriate discharge without coronary angiography. We posit that patients above the higher threshold (sensitivity 40.9%, specificity 80.8%) warrant at least stress testing or CT coronary angiography prior to discharge.

We further tested the use of sequential rather than single ECGs for prediction of revascularization and found that this approach significantly outperformed the use of individual ECGs. We posit that this paradigm should become standard practice in developing deep learning models using ECG data. In clinical practice, assessment of ECG evolution over time is a core component of ECG analysis, and it is intuitive that capturing intertemporal variation should improve a deep learning model’s performance. This principle also applies to other modalities of medical data, including imaging and lab data. Further research is warranted to study how such an algorithm would perform in clinical practice.

The influence of the PR and ST segments on AI-MI’s predictions is sensible but nevertheless interesting. While ST segment changes are well known markers of ACS, PR changes – including segment lengthening and depressions – tend to be more subtle and less commonly used. It is possible that AI-MI compares the relative height of PR and ST segments in making a prediction.

We found that a 1D ResNet model outperformed 1D Transformers and other more innovative models. EKG signals have unique characteristics that align well with CNN architectures: short-term temporal dependencies, consistent signal morphology within cardiac cycles, and local spatial correlations that are well-captured by convolutional filters. Transformers, on the other hand, excel in capturing long-range dependencies through attention mechanisms, but they carry high computational cost and lack the inherent inductive bias towards processing sequential signals with local correlations.

Key limitations of this study include the facts that 1) AI-MI was trained using data from a single center, 2) the model was validated using a held-out set from the same center, and 3) our analyses are retrospective and thus may not reflect AI-MI’s performance in clinical practice. In the future, it would be worthwhile to both train and validate AI-MI at clinical sites outside NYU Langone Health to ascertain its generalizability across health systems. Another key future direction would be to embed the model in an electronic health record and prospectively study its effect on clinician behavior and ACS outcomes. Finally, the sequential ECG approach should be applied to predict other outcomes of interest, including arrhythmias and cardiomyopathies.

## Conclusion

We developed AI-MI, a deep learning model utilizing sequential 12-lead ECG waveforms to predict whether a patient being worked up for ACS in the ED will undergo same-encounter coronary revascularization. AI-MI demonstrated high predictive performance, portending applicability for triaging potential ACS cases in the emergency department and for prioritizing patients for limited cardiac catheterization laboratory resources. The use of sequential ECG data significantly improved our model’s performance compared to using individual ECGs, reflecting the predictive value of temporal changes in ECG features over time. Grad-CAM analysis revealed that the PR and ST segments contributed most significantly to AI-MI’s decision-making process, aligning with clinical understanding of ST-segment changes as key indicators of myocardial ischemia.

## Data Availability

Electronic Health Record and ECG data used to train AI-ECG includes protected health information (PHI) from NYU Langone Health and cannot be made available. Raw data reflecting the performance of AI-ECG in various experiments will be made available on request.

## Clinical Perspectives

Differentiating acute coronary syndrome from low-risk chest pain etiologies is often nontrivial under the existing standard of care. However, early and accurate identification of acute coronary syndrome (ACS) is a key quality metric in Emergency Departments as diagnostic delay results in myocardial infarction, while unnecessary coronary angiography leads to adverse effects, prolonged hospital stays, and poor utilization of cardiac catheterization laboratory resources. AI- MI accurately predicts a patient’s need for same-encounter coronary revascularization during initial evaluation for anginal symptoms, portending potential use to risk stratify angina cases for early coronary angiography vs watchful waiting. In the future, AI-MI will be studied at NYU Langone in randomized clinical trials to evaluate its prospective efficacy in clinical practice.

