## Supplemental Information for "AI-MI: A Deep Learning Model to Predict Actionable Acute Coronary Syndrome Using 12-Lead ECGs"

**Supplementary Materials**

|  | **Predicted: Negative** | **Predicted: Indeterminate** | **Predicted: Positive** | **Total** |
| --- | --- | --- | --- | --- |
| **Actual: Negative** | 7704 | 3496 | 551 | 11751 |
| **Actual: Positive** | 28 | 258 | 303 | 589 |
| ***Percent Positive*** | *0.36* | *6.87* | *35.48* | *4.77* |

**Table S1:** Confusion matrix of AI-MI’s predictions for the “troponin ordered” cohort.

|  | **Predicted: Negative** | **Predicted: Positive** | **Total** |
| --- | --- | --- | --- |
| **Actual: Negative** | 9020 | 2731 | 11751 |
| **Actual: Positive** | 103 | 486 | 589 |
| ***Percent Positive*** | *1.13* | *15.11* | *4.77* |

**Table S2:** Confusion matrix of troponin assays for the “troponin ordered” cohort.

|  | **Predicted: Negative** | **Predicted: Indeterminate** | **Predicted: Positive** | **Total** |
| --- | --- | --- | --- | --- |
| **Actual: Negative** | 1207 | 877 | 647 | 2731 |
| **Actual: Positive** | 42 | 107 | 337 | 486 |
| ***Percent Positive*** | *3.36* | *10.87* | *34.25* | *15.11* |

**Table S3:** Confusion matrix of AI-MI’s predictions for the “troponin positive” cohort.

|  | **Predicted: Negative** | **Predicted: Indeterminate** | **Predicted: Positive** | **Total** |
| --- | --- | --- | --- | --- |
| **Actual: Negative** | 210 | 356 | 577 | 1143 |
| **Actual: Positive** | 47 | 193 | 589 | 829 |
| ***Percent Positive*** | *18.29* | *35.15* | *50.51* | *42.04* |

**Table S4:** Confusion matrix of AI-MI’s predictions for the “coronary angiogram” cohort.

 
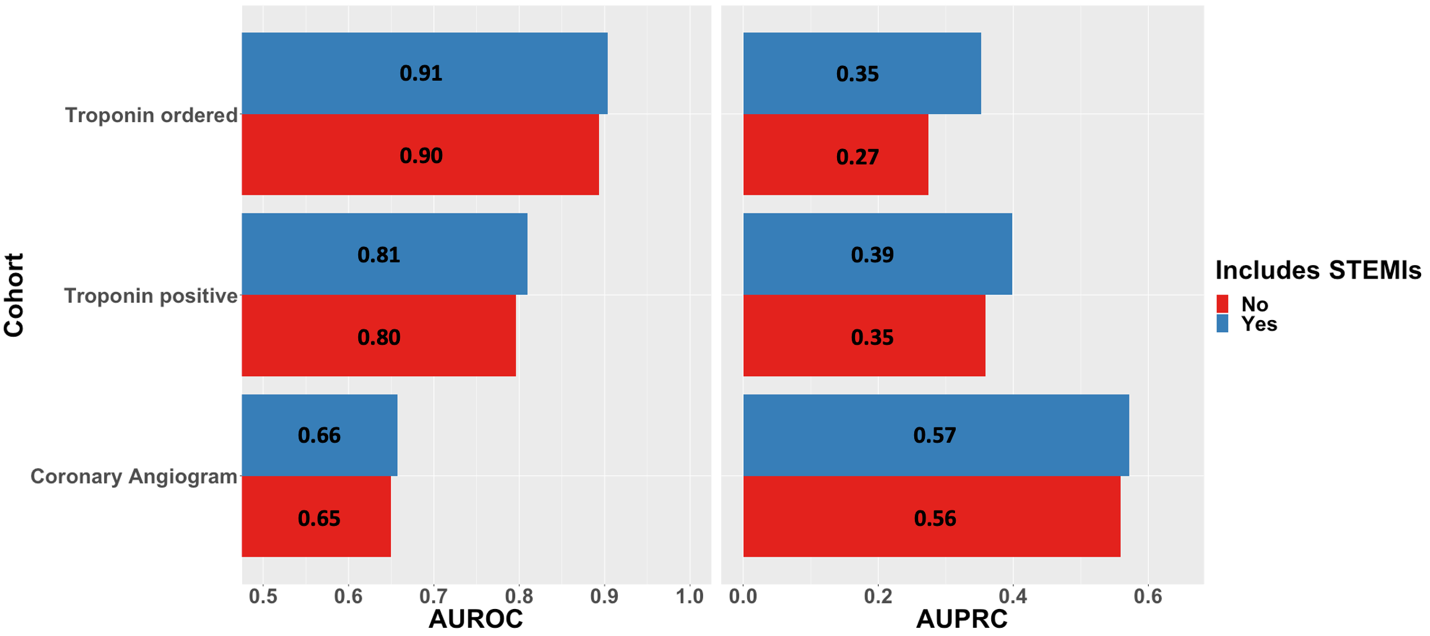

**Figure S1:** AUROCs and AUPRCs of AI-MI on cohort including (blue) and not including (red) patients flagged by NYU’s automated EHR reader as STEMIs. This suggests that AI-MI’s performance in this study is not simply driven by successfully identification of STEMIs.

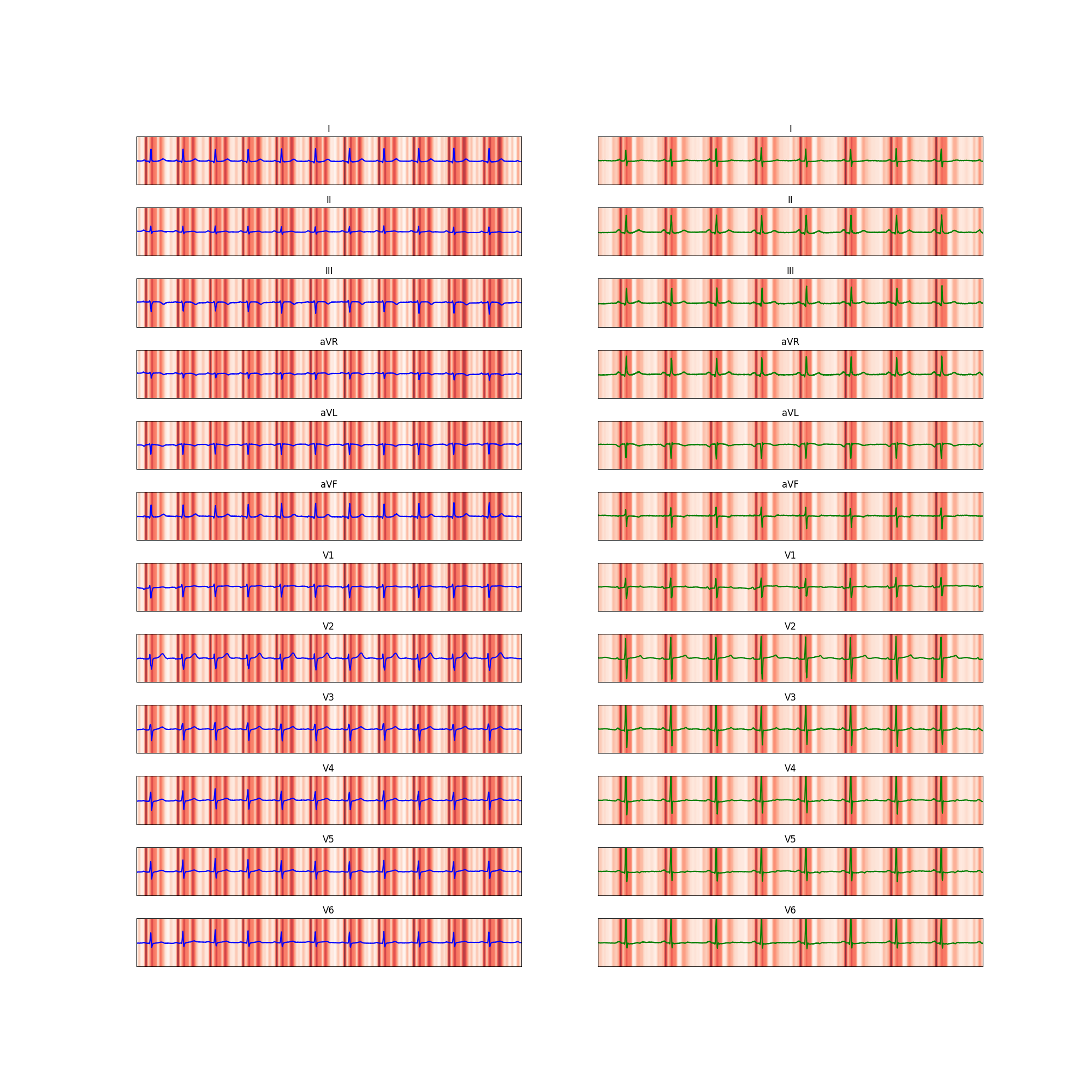
**(i)**
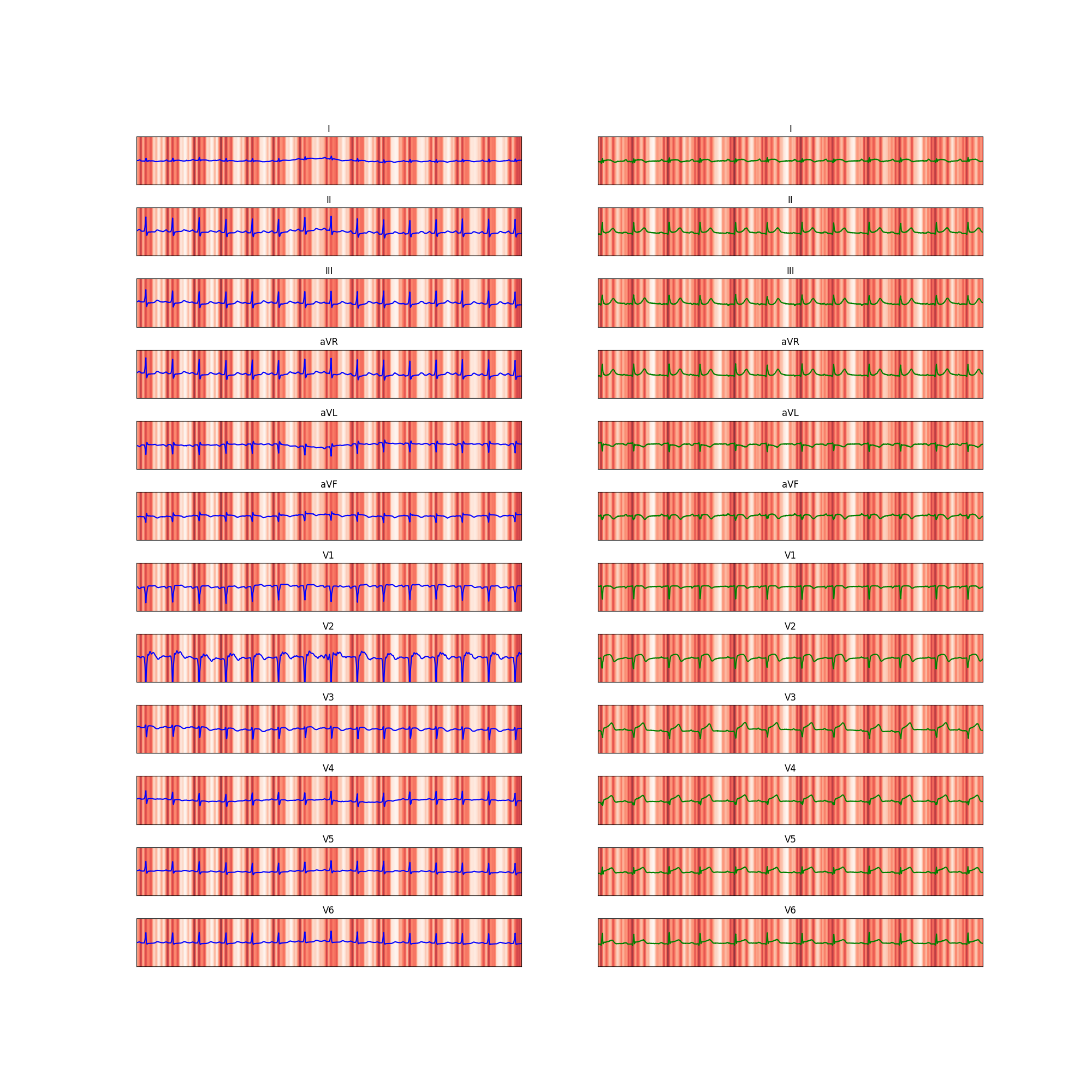
**(ii)**
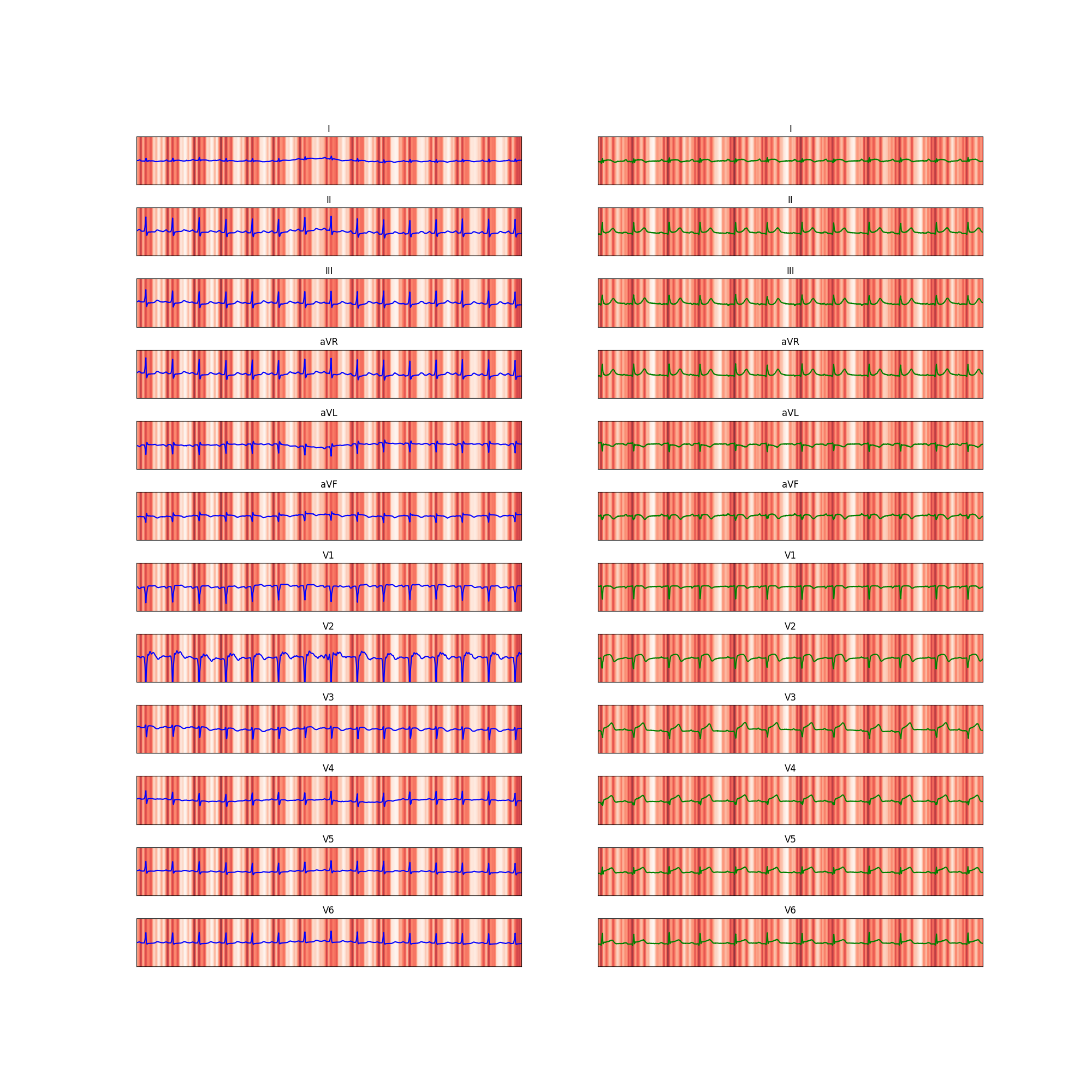
**(iii)**
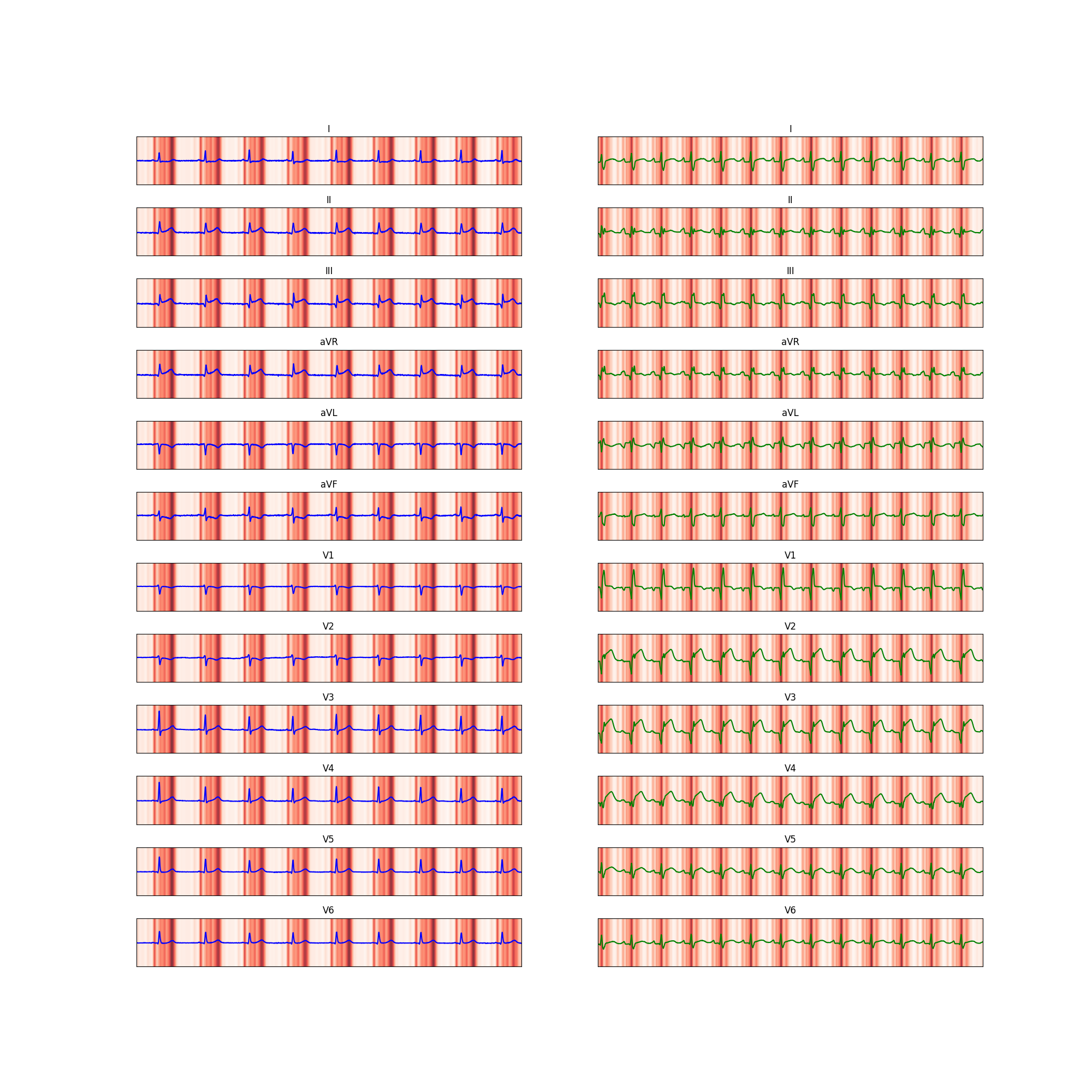
**(iv)**
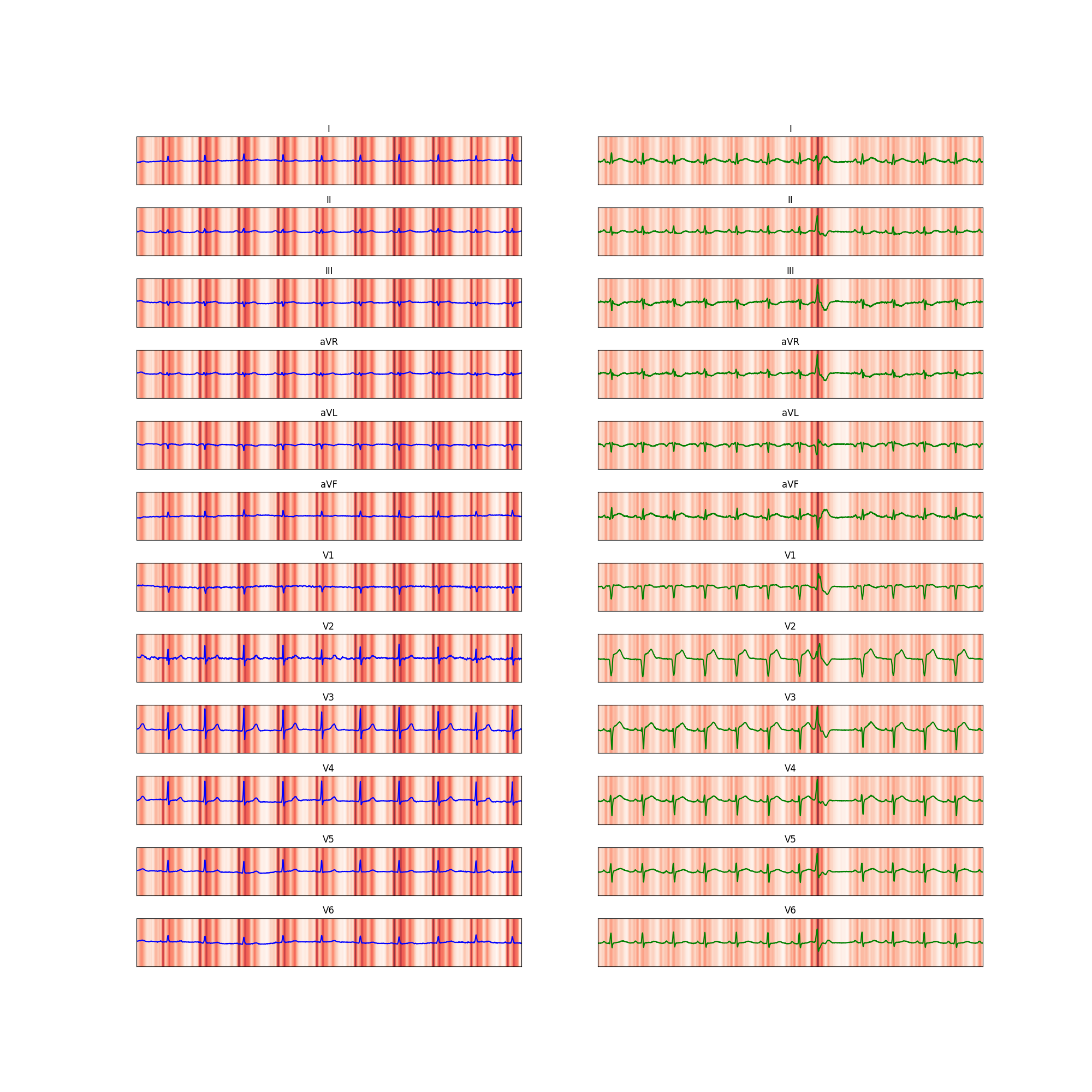
**(v)**
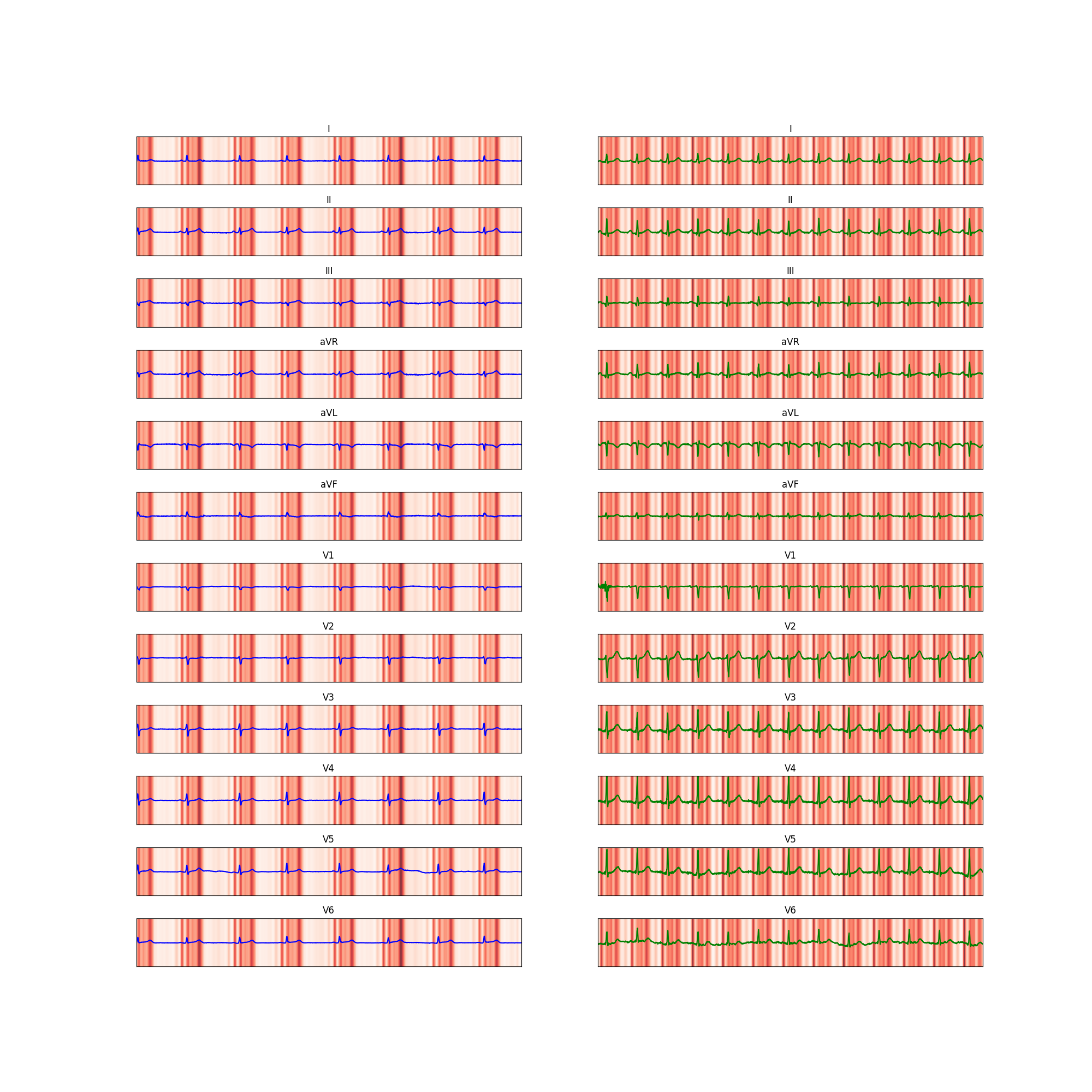
**(vi)**
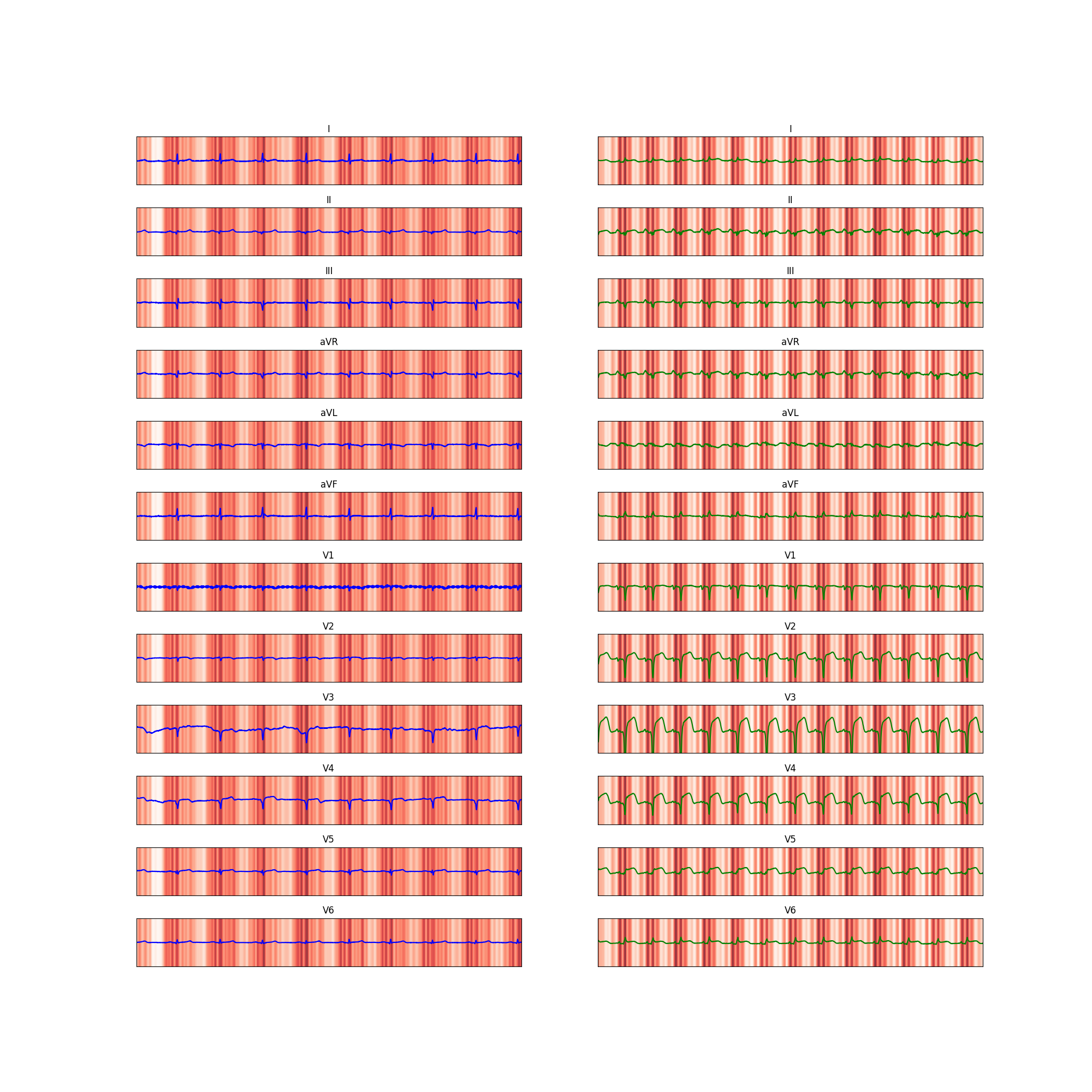
**(vii)**
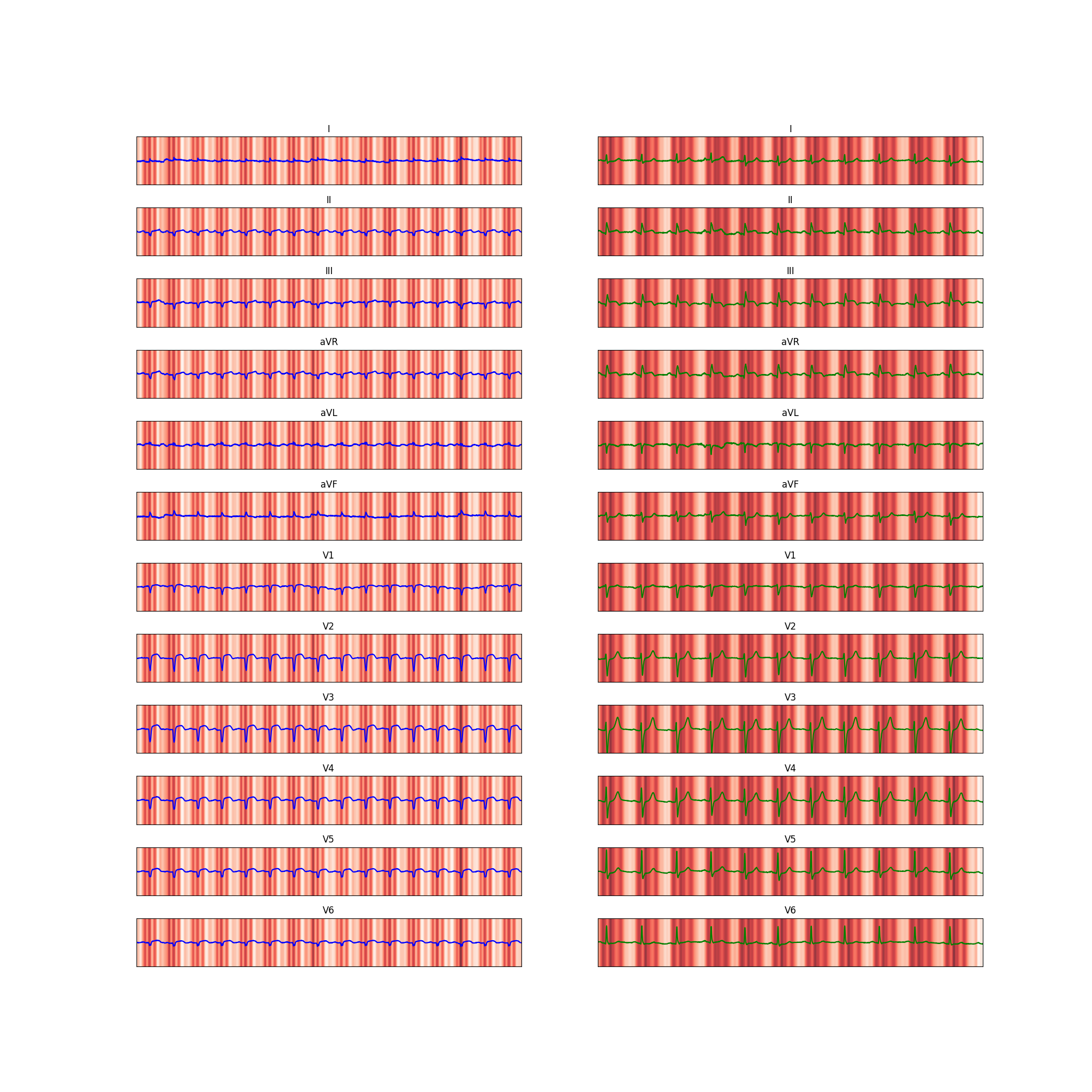
**(viii)**
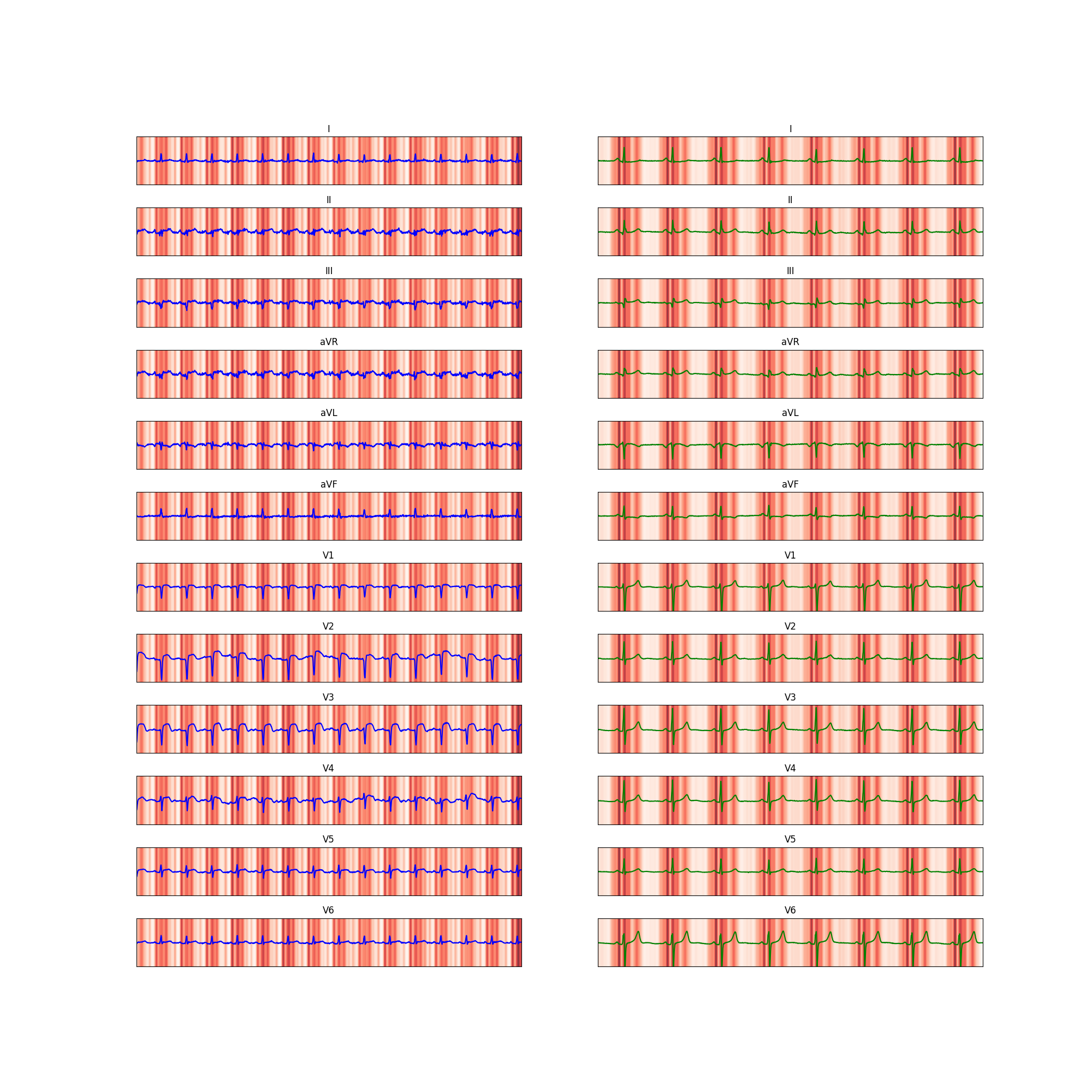
**(ix)**

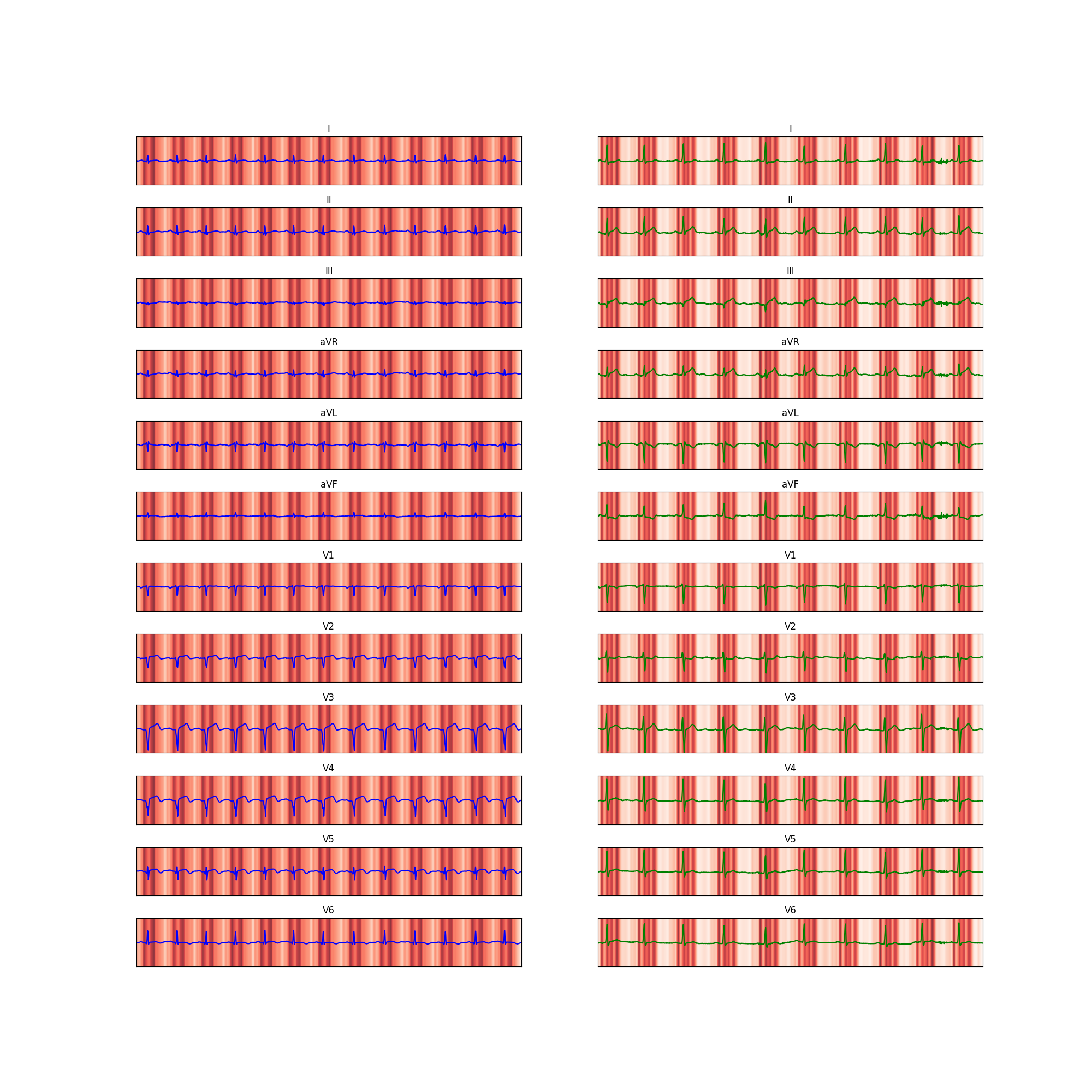
**(x)**

**Figure S2:** Grad-CAM heatmaps displaying regions of ECGs driving our model’s predictions for (left) the patient with ACS for whom our model predicted the second (ii) through tenth (x) highest probabilities of ACS (i.e. top true positives), and (right) the patient without ACS for whom our model predicted the second (ii) through tenth (x) highest probabilities of ACS (i.e. top false positives). Heatmaps were generated using the Grad-CAM algorithm. Corresponds to Figure 4 in the main text.

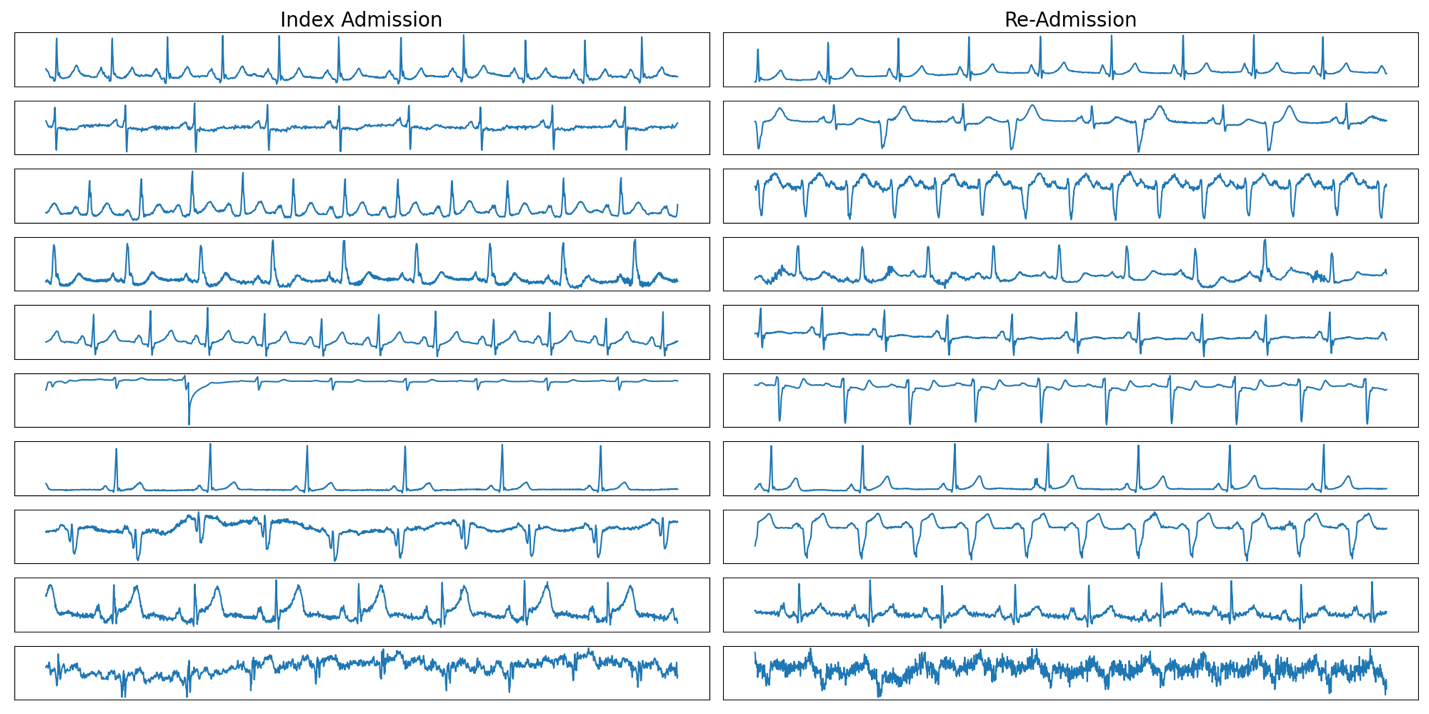

**Figure S3:** Initial EKG lead IIs of patients who were predicted by AI-MI to be high-risk for same-encounter revascularization, were discharged, and were subsequently re-admitted for urgent re-vascularization within 1 month. Waveforms are shown during index admission (left) and at readmission (right).
